# Clinical Testing of Pediatric Mpox Specimens: Unique Features and Challenges in a Low Prevalence Population

**DOI:** 10.1101/2023.01.20.23284754

**Authors:** Angela Ma, Benjamin T. Bradley

## Abstract

**BACKGROUND:** Pediatric mpox cases comprise less than 0.3% of the total cases reported in the United States during the global 2022 outbreak. As a result, relatively little is known about the epidemiology or performance characteristics of clinical testing in this group.

**METHODS:** We retrospectively extracted and analyzed results for pediatric mpox specimens tested at a national reference laboratory from July to December 2022.

**RESULTS:** During our study period 13.4% (2,063/15,385) of specimens were from individuals <18 years of age. The positivity rate of pediatric specimens was significantly lower than in adults (1.3% vs 22.3%). The pediatric cohort also consisted of a higher percentage of females (42.7% vs 31.0%) and lower percentage of specimens from genital sources (9.0% vs 19.7%) as compared to adults. In children, specimens were most frequently collected from 1-year-olds (10.1%) and least frequently from 11-year-olds (3.5%). Positivity rates were disproportionately elevated in the less than 1-year and 17-year-old age groups (7.8% and 6.4%, respectively). Ct values of positive cases were not statistically different between pediatric and adult cohorts (25.2 vs 22.2, p>0.05). When all pediatric cases with an initial positive mpox result were examined, 5/26 were classified as inconclusive and 2/26 were determined to be false positives. Based on these results, the positive predictive value of monkeypox virus detection was 90.5% (95% CI: 70.4-97.4%) in children.

**CONCLUSION:** These results highlight important differences between pediatric and adult mpox populations and reinforce the need for clinical correlation when reporting positive results from a low prevalence population.

## Introduction

Monkeypox virus (MPXV) is the etiologic agent of mpox (1). Historically, mpox has been observed in endemic regions of Africa with rare, sporadic outbreaks in non-endemic regions (2, 3). However, in the spring of 2022, several non-endemic countries began reporting sustained human-to-human transmission of MPXV (4). The resulting global outbreak has led to mpox being declared a Public Health Emergency of International Concern by the World Health Organization (5). Epidemiological reports of the 2022 mpox outbreak have differed from prior accounts of mpox in several important ways. Historically, children have been most commonly affected and present with lesions on the extremities (6). The current outbreak has predominately affected populations of men who have sex with men and has been associated with transmission during intimate contact (7). The Centers for Disease Control and Prevention estimate less than 0.3% of mpox cases occur in individuals less than 18 years of age (8).

Traditionally, children have comprised the largest group affected by mpox with the median age of infection being 4 and 5 years old for cases reported in the 1970s and 1980s; however, the median age has increased in recent decades (6). Childhood mpox has also been associated with a high mortality rate, up to 10% during the 1980s (9). However, in a recent meta-analysis of Clade II MPXV cases prior to 2022, 22 pediatric cases were identified with only one documented death (10). During the current outbreak, reports from Spain and the United States found severe complications were rare in pediatric patients and no intensive care unit admissions or deaths were documented (8, 11). For cases of mpox in children less than 18 years old, two main transmission routes have been identified, each specific to certain age groups. For individuals less than 13 years of age, transmission from household contacts the most common source of infection. Whereas, in adolescents, sexual activity was the source for 97% of infections with a known transmission route (8).

Diagnosing infectious diseases in a low-risk population presents a challenge to clinical microbiology laboratories. The most concerning dilemma is the risk for reporting false positive results due to poor positive predictive value in a low prevalence setting. As demonstrated during the SARS-CoV-2 pandemic, the clinical value of a positive result was compromised when surveillance testing was performed in a low-risk population (12). Mpox testing in pediatric populations is also complicated by the presence of viral exanthems which may present with similar clinical features including varicella, molluscum contagiosum, and hand foot and mouth disease (13). False positive results may lead to inappropriate treatment, unnecessary prophylactic vaccination of close contacts, and unintended social stigma (14). To better understand how mpox testing differs from adult populations and examine relevant differences in positivity rates, we performed a retrospective review of all pediatric mpox testing performed at a national reference laboratory between July 25^th^ and December 14^th^, 2022.

## Methods

### Ethics Statement

An IRB exemption was granted by the University of Utah IRB under ID#: 00158025.

### MPXV NAAT Testing

Detection of MPXV DNA was performed as previously described (15). In brief, specimens were either extracted on the Chemagic MSMI instrument (PerkinElmer) followed by amplification on the QuantStudio 12K Flex (ThermoFisher) or extraction and amplification was performed on the cobas 6800 platform (Roche Diagnostics). Both assays used lab-developed primers and probes which targeted a conserved region of the viral polymerase gene common to the Orthopoxvirus family (16).

### Data Analysis and Statistics

Inhibited and invalid results, comprising 0.04% of results, were excluded from analysis. Specimens were classified as derived from genital sites if they contained any of the following words: “anal”, “buttock”, “gluteal”, “groin”, “inguinal”, “labia”, “penis”, “perineum”, “pubic”, “rectal”, “scrotum”, “vagina”, “vulva” or variations of such words. Deidentified data were analyzed in Prism Graphpad v. 9.4.1. Statistical tests included Chi-squared, Mann-Whitney, Fisher’s exact, and Welch’s t-test. The cutoff for statistical significance for all tests was established as 0.05.

Specimens were categorized as “detected”, “inconclusive”, or “not detected”. Specimens with no repeat testing were categorized as follows: detected if Ct<34, inconclusive if Ct=34-50, and not detected if CT >50. Specimens with repeat testing were categorized as follows: detected if Ct <50 for both replicates, inconclusive if one replicate with Ct<50 and the other Ct>50, and not detected if both replicates with Ct>50.

Patient cases were classified as “positive”, “inconclusive”, or “false positive”. A positive case was classified as any individual with at least one detected specimen. An inconclusive case was classified as any individual with at least one inconclusive specimen and no detected specimens. Cases were classified as false positive only in situations where the ordering provider provided a clinical history not consistent with mpox and repeat testing of the specimen was not detected.

## Results

### Pediatric mpox specimens have different demographics and positivity rates from adult specimens

During the study period, pediatric specimens comprised 13.4% (2,063/15,385) of the total mpox testing volume (Fig 1A). The proportion of pediatric to adult individuals within the cohort was similar at 13.6% (1,480/10,862). The pediatric cohort was comprised of 56.8% male, 42.7% female, and 0.5% of individuals with unknown sex, while the adult cohort contained 66.8% male, 31.0% female, and 2.2% of individuals with unknown sex (Fig 1B). The sex distribution was significantly different between the two age groups (p<0.001, chi-squared test, individuals with unknown sex excluded). 9.0% of specimens were collected from genital sources in the pediatric cohort while 19.6% of adult specimens were collected from genital sources (p<0.001, chi-squared) (Fig 1C). The positivity rates of the pediatric and adult cohorts were 1.3% and 22.3%, respectively. When stratified by sex, the positivity rate of the female pediatric cohort was 1.1% and the male pediatric cohort was 1.4% (p=0.565, Fisher’s exact test). In the adult cohort, the positivity rate between females and males was 3.2% and 31.1% (p<0.0001, chi-squared) (Fig 1D). The median Ct value of specimens from pediatric individuals positive for mpox was 25.2 while the median Ct value for adult specimens was 22.2. The difference was not statistically significant (p=0.146, Mann-Whitney test) (Fig 1E).

**Figure 1.**
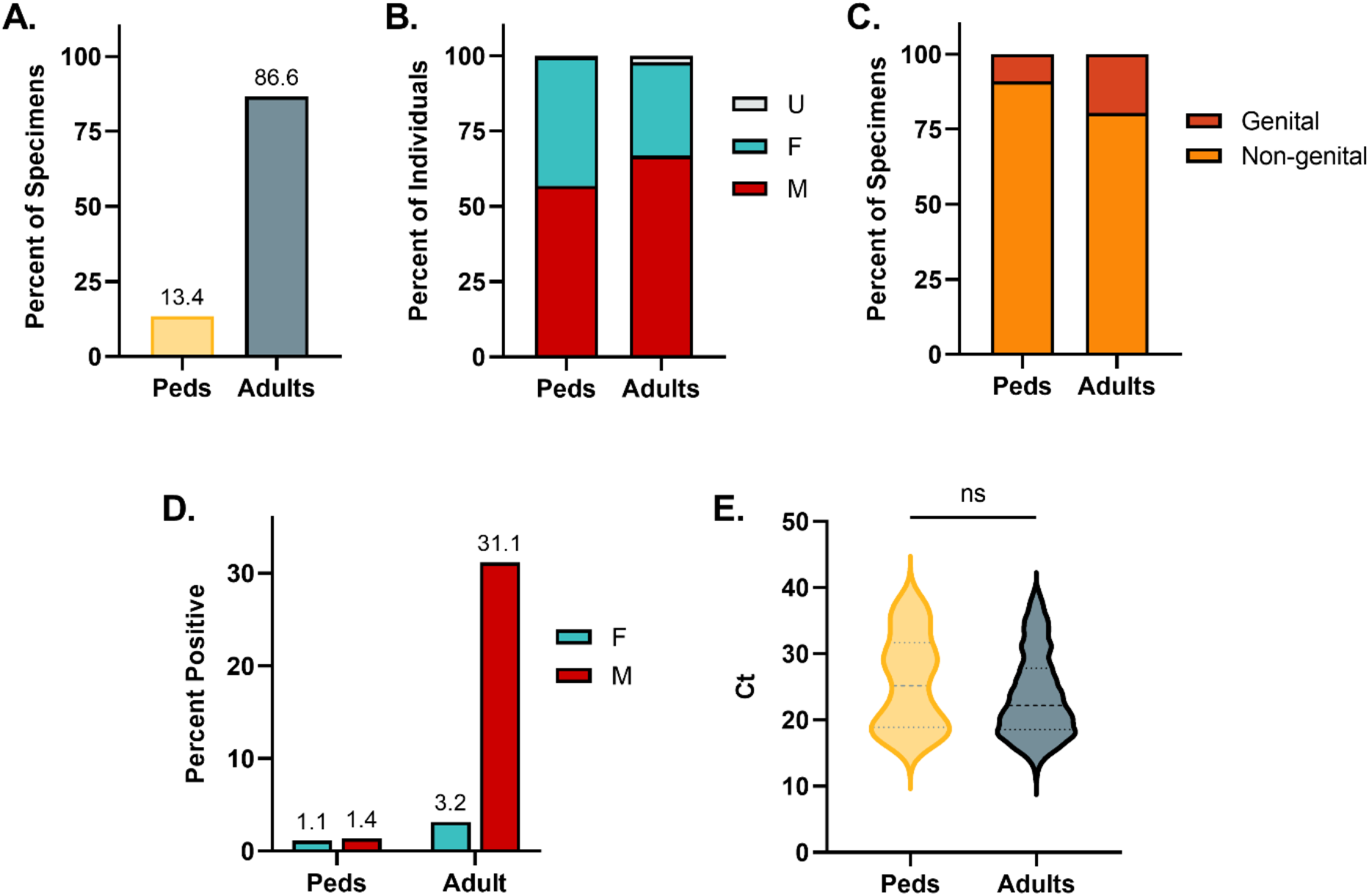
Demographic and Clinical Data for Mpox Specimens from Pediatric and Adult Individuals. (A) Percent of total specimen volume. (B) Percent of female, male, and unknown sex. (C) Percent of specimens obtained from genital or non-genital sources. (D) Positivity rate of pediatric and adult specimens stratified by sex. (E) Comparison of Ct values between pediatric and adult specimens positive for mpox

### Mpox positivity rates are highest at the extremes of the pediatric age group

The median age and standard deviation of the pediatric cohort was 6 ± 5.4 years. 1-year-olds were the most frequently tested group (10.1%), followed by 2-year-olds (9.2%), 3-year-olds (8.2%), and 17-year-olds (7.3%). 11-year-olds were the least frequently tested group (3.5%) (Fig 2A). Age distribution of mpox cases demonstrated bimodal peaks with the highest number of positive cases in 17-year-olds (36.8%) followed by <1-year-olds (26.3%). Single positive cases were identified in 1, 2, 6, 8, 9, 14, and 16 year-olds (Fig 2B). Mpox positivity rates by age were calculated as follows: <1 year of age – 7.8%, 1-year-olds – 0.9%, 2-year-olds – 0.7%, 6-year-olds – 1.1%, 8-year-olds - 1.4%, 9-year-olds – 1.9%, 14-year-olds – 1.6%, 16-year-olds – 1.3%, and 17-year-olds – 6.4%. The mean age for pediatric mpox diagnosis was significantly earlier for females (4.5 years) than males (11.6 years) (p=0.039, Welch’s t test). Ct values for positive cases ranged from 18.2 to 37.1 based on the earliest Ct available per patient. Ct values were not statistically different between age groups or sex (Fig 2C).

**Figure 2.**
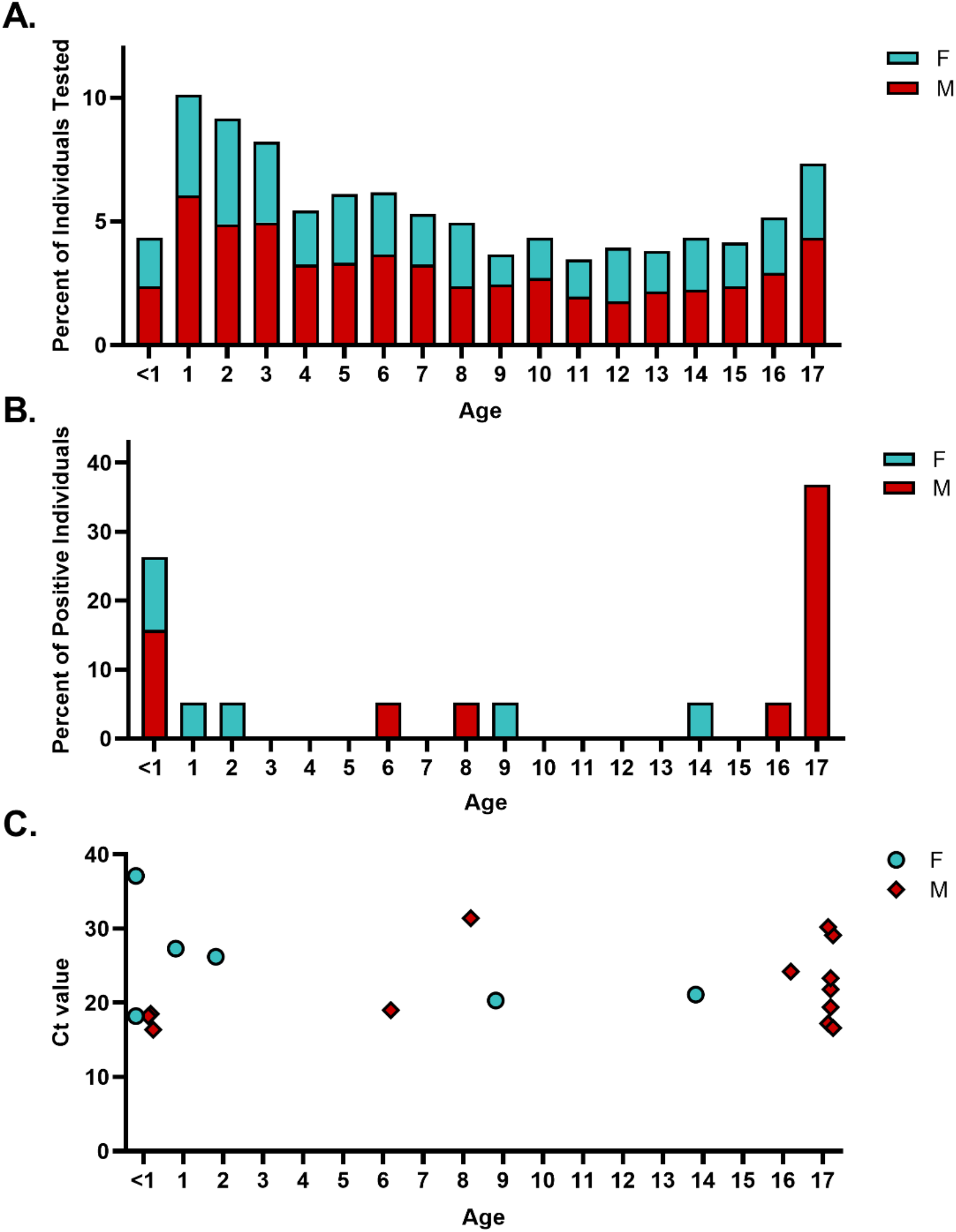
Mpox Results by Age and Sex in the Pediatric Cohort. (A) Age distribution of total pediatric mpox testing. (B) Age distribution of positive mpox cases. (C) Earliest Ct value for individual positive cases of mpox.

### A significant proportion of mpox detected in children result in inconclusive or false positives results

Of the 26 individuals with MPXV amplified at least once from a specimen, 19 (73%) were determined to be positive for mpox, 5 (19%) inconclusive for mpox, and 2 (8%) false positive for mpox (Table 1). Excluding the inconclusive specimens, assay sensitivity and specificity was 100% (95% CI: 82.4-100%) and 99.9 (95% CI: 99.5-100.0), respectively. The positive predictive value was 90.5% (95% CI: 70.4-97.4%) and the negative predictive value was 100% (no known false negative cases).

**Table 1.**
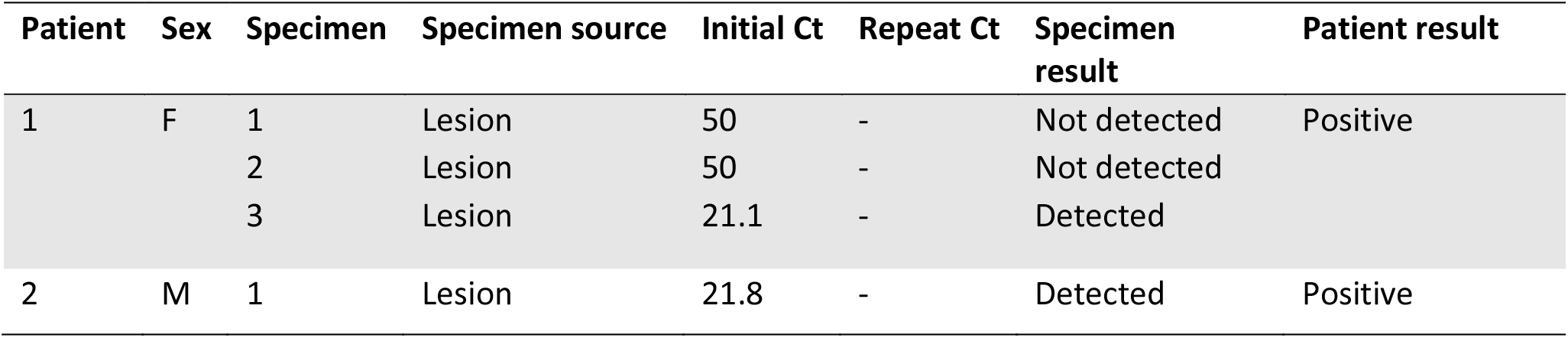
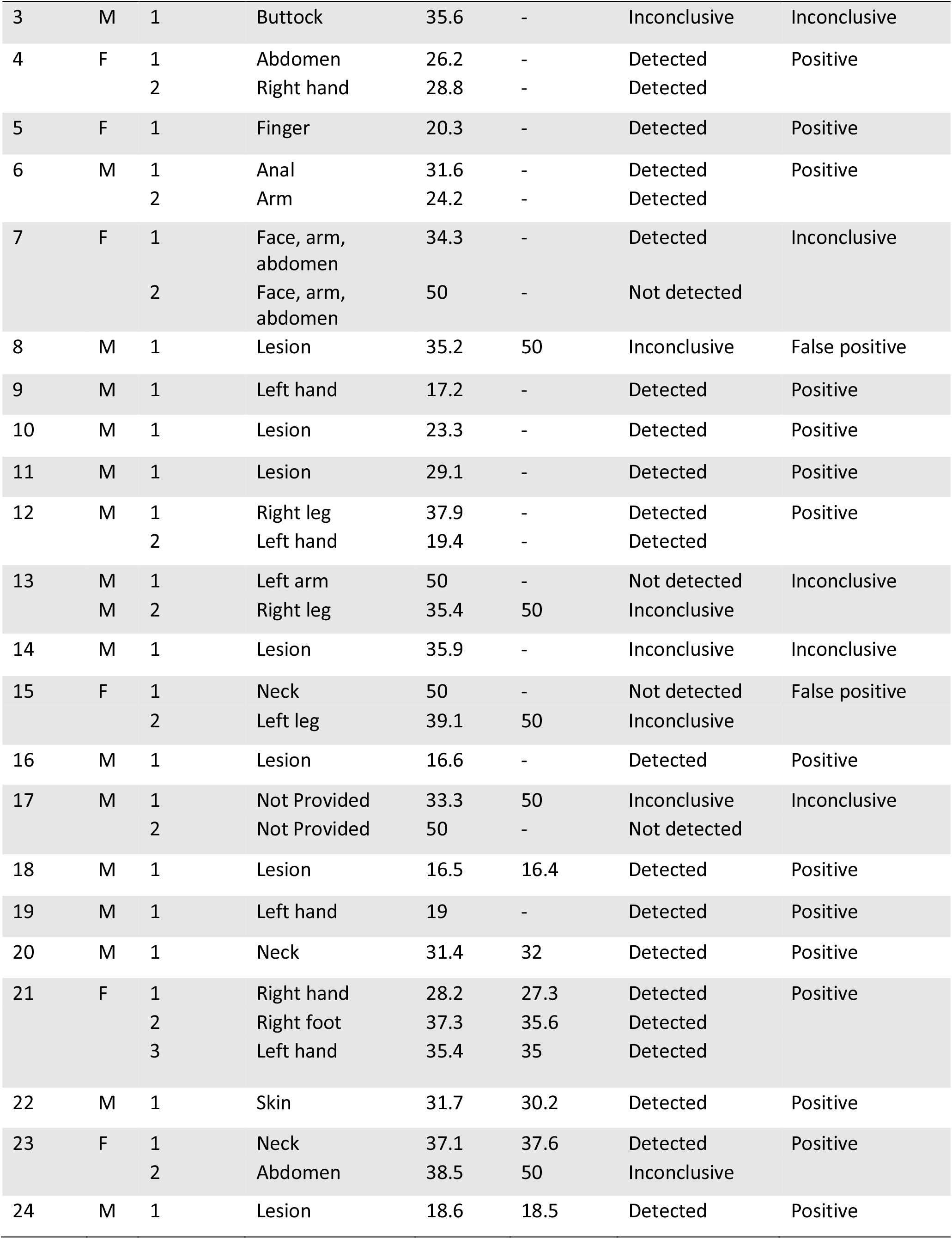
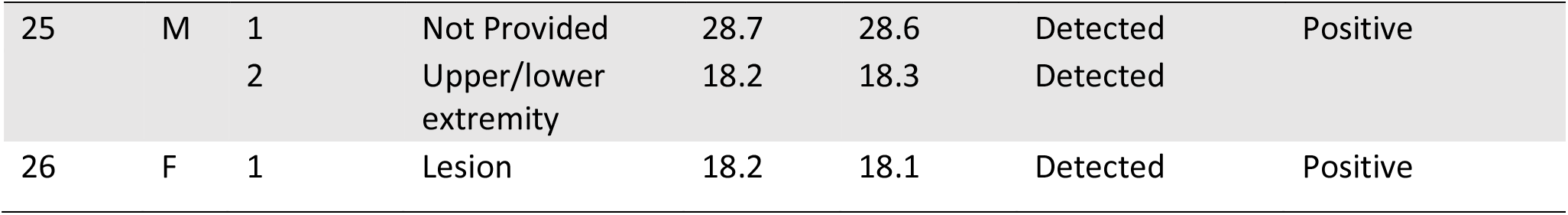
Pediatric specimens with mpox signal detected.

| Patient | Sex | Specimen | Specimen source | Initial Ct | Repeat Ct | Specimen result | Patient result |
| --- | --- | --- | --- | --- | --- | --- | --- |
| 1 | F | 1 | Lesion | 50 | - | Not detected | Positive |
|  |  | 2 | Lesion | 50 | - | Not detected |  |
|  |  | 3 | Lesion | 21.1 | - | Detected |  |
| 2 | M | 1 | Lesion | 21.8 | - | Detected | Positive |
| 3 | M | 1 | Buttock | 35.6 | - | Inconclusive | Inconclusive |
| 4 | F | 1 | Abdomen | 26.2 | - | Detected | Positive |
|  |  | 2 | Right hand | 28.8 | - | Detected |  |
| 5 | F | 1 | Finger | 20.3 | - | Detected | Positive |
| 6 | M | 1 | Anal | 31.6 | - | Detected | Positive |
|  |  | 2 | Arm | 24.2 | - | Detected |  |
| 7 | F | 1 | Face, arm, abdomen | 34.3 | - | Detected | Inconclusive |
|  |  | 2 | Face, arm, abdomen | 50 | - | Not detected |  |
| 8 | M | 1 | Lesion | 35.2 | 50 | Inconclusive | False positive |
| 9 | M | 1 | Left hand | 17.2 | - | Detected | Positive |
| 10 | M | 1 | Lesion | 23.3 | - | Detected | Positive |
| 11 | M | 1 | Lesion | 29.1 | - | Detected | Positive |
| 12 | M | 1 | Right leg | 37.9 | - | Detected | Positive |
|  |  | 2 | Left hand | 19.4 | - | Detected |  |
| 13 | M | 1 | Left arm | 50 | - | Not detected | Inconclusive |
|  | M | 2 | Right leg | 35.4 | 50 | Inconclusive |  |
| 14 | M | 1 | Lesion | 35.9 | - | Inconclusive | Inconclusive |
| 15 | F | 1 | Neck | 50 | - | Not detected | False positive |
|  |  | 2 | Left leg | 39.1 | 50 | Inconclusive |  |
| 16 | M | 1 | Lesion | 16.6 | - | Detected | Positive |
| 17 | M | 1 | Not Provided | 33.3 | 50 | Inconclusive | Inconclusive |
|  |  | 2 | Not Provided | 50 | - | Not detected |  |
| 18 | M | 1 | Lesion | 16.5 | 16.4 | Detected | Positive |
| 19 | M | 1 | Left hand | 19 | - | Detected | Positive |
| 20 | M | 1 | Neck | 31.4 | 32 | Detected | Positive |
| 21 | F | 1 | Right hand | 28.2 | 27.3 | Detected | Positive |
|  |  | 2 | Right foot | 37.3 | 35.6 | Detected |  |
|  |  | 3 | Left hand | 35.4 | 35 | Detected |  |
| 22 | M | 1 | Skin | 31.7 | 30.2 | Detected | Positive |
| 23 | F | 1 | Neck | 37.1 | 37.6 | Detected | Positive |
|  |  | 2 | Abdomen | 38.5 | 50 | Inconclusive |  |
| 24 | M | 1 | Lesion | 18.6 | 18.5 | Detected | Positive |
| 25 | M | 1 | Not Provided | 28.7 | 28.6 | Detected | Positive |
|  |  | 2 | Upper/lower extremity | 18.2 | 18.3 | Detected |  |
| 26 | F | 1 | Lesion | 18.2 | 18.1 | Detected | Positive |

Based on additional clinical information obtained following discussion with the ordering provider, two cases (Pt 8 and Pt 15) were determined to represent false positive results. Both of these cases demonstrated late Ct values of 35.2 and 39.1 that did not repeat following re-extraction and amplification. Additionally, one case was adjudicated as positive despite one of two specimens not repeating. This patient (Pt 23) initially had late Cts detected in both specimens (37.1 and 38.5). The 37.1 specimen repeated at 37.6, however, the 38.5 specimen did not repeat.

When positive samples were repeated (n=16), the median Ct value shifted by 0.1 cycles (range: - 1.7 to 0.6 cycles). For samples that failed to repeat, the earliest Ct value that did not yield a repeatable result was 33.3 (Pt. 17). Repeat testing of all positive pediatric results was instituted during the study period on September 17^th^, 2022. During that time 11/12 samples in which MPXV was initially detected repeated as positive. The specimen that did not repeat was from the previously described patient (Pt 23).

## Discussion

Recently, more clinical data is available describing the manifestations of mpox in the pediatric population (8, 11, 17–19). However, this information is mostly outside the context of laboratory testing. In this study we examined the demographics and results of pediatric mpox testing at a large reference laboratory. The results of our study reinforce clinical trends that have been identified and add further clarity on higher risk groups within the pediatric population.

The relative percent of pediatric specimens tested in our laboratory decreased from 14.1% when examined in September 2022 to 13.4% by December 2022 (15). Epidemiologic studies of mpox have found a higher relative percentage of female cases among children (20%-36%) than adults and noted an absence of genital lesions in those less than 13-years old (8, 11). This is reflected in the ordering practices for pediatric mpox specimens in our study. We found a higher number of female individuals (42.7% vs 31.0%) and non-genital specimens (91% vs 80.4%) in children versus adults.

When examining the distribution of age and gender for pediatric mpox specimens, several trends were observed. First in terms of overall testing volume, 1-to 3-year-olds were the most frequently tested groups with 11-year-olds the least frequently tested. It is interesting that those less than 1 year old were not tested at a higher volume, given over 25% of cases occurred in this age group. This may suggest other types of close contact outside of interactions with caregivers led to preferential testing in 1-to 3-year-olds. Potential exposures may include attendance of community events which was identified as a risk factor for childhood HSV-1 acquisition (20). Second, positivity rates were strongly polarized to the less than 1-year and 17-year-old age groups. The less than 1 age group was comprised of female and male cases while the 17-year-old group was exclusively male. These findings reflect the different modes of transmission between age groups, with infants more likely infected by close skin-to-skin contact from caregivers and adolescents infected through sexual contact (8). Ct values between the less than 1- and 17-year-old age groups were similar, suggesting that route of infection does not play a significant role in viral replication. When the positivity rates of individual age groups were compared to adults, both the less than 1- and 17-year-old age groups demonstrated higher positive rates than adult females. Increased surveillance for mpox in these age groups may be warranted in future outbreaks.

There are at least two documented cases of false positive mpox results from pediatric specimens with late Ct values (14). To address this issue, the CDC recommended samples with a Ct of 34 or greater undergo repeat testing (21). Following this recommendation, our institution began automatically repeating all positive samples from patients <18 years old. Prior to this, repeat testing was performed based on clinical request or medical director discretion. The majority of specimens which did not repeat (4 of 5) in our study were greater than 34 cycles. However, there was one specimen (Pt 17, specimen 1) with an earlier Ct value of 33.3 which did not repeat. The second sample from this patient was negative, and so the case was classified as inconclusive. If following the CDC guidelines, the first specimen would not have been sent for repeat testing and may have led to a false positive result.

Failure of a late Ct to repeat should not be used as the sole factor to report a specimen as positive or negative. True cases of mpox may also have samples that fail to repeat due to late Cts. For example, Pt 23 had two specimens with late Cts (37.1 and 38.5) with the second specimen failing to repeat. Given the similar presence of low MPXV in both specimens, we hypothesized these findings are more reflective of an individual late in the disease course. Failure to re-amplify the 38.5 Ct specimen was likely due to nucleic acid present below the limit of detection. Because Ct values of laboratory developed tests can be influenced by a myriad of factors (primer/probe design, cycling conditions, curve analysis), it is imperative that labs performing mpox testing have a robust understanding of their assay and criteria for repeating. When a patient has discordant results or a late Ct result, it is reasonable for the laboratory to repeat extraction and amplification of the specimens. Additionally, consultation with the ordering provider may provide helpful context to the patient’s exposure risks. However, even when clinical information is available, epidemiological investigations may not provide a conclusive route of mpox exposure (18). In a series of 83 cases of pediatric mpox in the US, 34% (28/83) were from unknown routes of transmission (8).

Given the descriptive nature of our study and incomplete patient histories, our conclusions have several limitations. In our role as a reference laboratory, we lacked clinical information for the majority of pediatric cases where mpox was detected. As a result, we retrospectively classified cases as “positive” or “inconclusive” based on laboratory results and Bayesian reasoning. It is highly likely some of the inconclusive cases are false positives and would further lower the positive predictive value of mpox detection. Our guidelines for repeat testing of pediatric specimens changed over the course of the study, and there were 4 samples with late Cts that were not repeated.

In summary, the pediatric age group comprises a minority of those tested for mpox and the overall positivity rate is much lower than in adult males. Specimens from children are more frequently from females and collected from non-genital sources. The age groups at highest risk of mpox appear to be those less than 1 year of age and those 17 years of age. Inconclusive and false positive results comprise a significant number of cases in which mpox is initially detected. Careful clinical correlation and repeat testing of patient samples should be considered within this age group.

## Data Availability

All data produced in the present study are available upon reasonable request to the authors.

